# Introduction to and spread of COVID-19 in care homes in Norfolk, UK

**DOI:** 10.1101/2020.06.17.20133629

**Authors:** Julii Brainard, Steven Rushton, Tim Winters, Paul R. Hunter

**Affiliations:** Norwich Medical School, University of East Anglia Norwich NR4 7TJ, UK; School of Natural and Environmental Sciences, Newcastle University; Norfolk County Council, Norfolk NR1 2DH UK

**Keywords:** Personal protection equipment, COVID-19, care homes, Cox-proportional hazards model, mixed effect models

## Abstract

**BACKGROUND:** Residential care homes for the elderly have been important settings for transmission of the SARS-CoV-2 virus that causes COVID-19 disease. METHODS: We undertook a secondary analysis of a dataset about 248 care homes in the county of Norfolk, eastern England. The dataset recorded categories of staff (nurses, care workers and non-care workers), their status (available, absent due to leave or sickness and extra staff needed to address the coronavirus pandemic) in the period 6 April −6 May 2020. Counts of residents (if any) at each care home with COVID-19 were also available, as well as descriptions of access by the home to personal protection equipment (PPE: gloves, masks, eye protection, aprons and Sanitiser). PPE access was categorised as (most to least) green, amber or red. We undertook two stage modelling, first for any detection of COVID-19 in the homes, and a second model to relate any increases in case counts after introduction to staffing or PPE levels. RESULTS: We found that the counts of non-care workers had strongest relationships (and only link significant at p < 0.05) to any introduction of SARS-CoV-2 to the homes. After a home had at least one detected case, higher staff levels and more severe PPE shortages were most linked to higher case counts (p < 0.05) during the monitoring period. CONCLUSION: Better managing aspects of staff interaction with residents and some working practices should help reduce ingression to and spread of COVID-19 within residential homes for the elderly.

**THUMBNAIL SKETCH:** What is already known on this subject?

➢ Close to 40% of all UK with COVID-19 deaths in early May of 2020 were among care home residents.
➢ That this care sector was underfunded and under-equipped to prevent disease introduction and spread is recognised but the mechanisms of how disease entered or spread have not been quantified.

What this study adds?

➢ Detection of any COVID-19 cases in homes was directly linked to the counts of staff members who were not directly involved in personal care.
➢ Subsequent disease spread was directly most strongly linked to lack of facemasks and eye protection, somewhat less to total counts of care workers employed.
➢ The findings demonstrate an inverse strong link between available PPE and case counts in care homes after disease became present.

## INTRODUCTION

Residential care homes for the elderly have been important settings for transmission of the SARS-CoV-2 virus that causes COVID-19 disease [1]. It became apparent early in the COVID-19 pandemic that infection control within care homes for the elderly would be especially challenging and yet important to reducing total mortality and wider disease spread. The viral disease has disproportionately more severe outcomes among the elderly [2] while residential settings are well understood to be places where any disease outbreak can be especially difficult to control. That care homes for the elderly were key foci for transmission was quickly recognised in many countries and territories, including places that nominally were otherwise able to quite effectively contain and control spread of SARS-CoV-2 [3]. The high level of COVID-19 deaths in care home settings became highly politicised [4-7].

Challenges in preventing or controlling infectious disease outbreaks in care settings are myriad. In many countries, the elderly care sector is acknowledged to be under-funded and staffed by relatively low-paid workers who may have insufficient training or experience in infection control [7, 8]. A detailed outbreak report on a COVID-19 outbreak in February-March 2020 focused on the Kirkland Care facility in Washington State, USA [9]. This nursing home provided care for over 100 residents. Many factors were identified that contributed to late recognition and failure to prevent SARS-CoV-2 infection in many dozens of persons linked to this outbreak, who were mostly residents, some staff and at least one visitor. To recap, the most important vulnerability factors identified in the Kirkland outbreak were (in no particular order):

- Delayed awareness of the disease (“low index of suspicion”)
- Staff who worked while symptomatic
- Staff who worked in multiple care facilities (increasing their own personal risk of exposure and facilitating disease transfer between institutions)
- Inadequate familiarity with and adherence to PPE recommendations
- Inadequate supplies of PPE and hand sanitizer
- Limited availability of testing
- Difficulty identifying symptomatic residents

Within the UK, personal protection equipment (PPE, such as masks, gloves and protective gowns) for care home workers became publicly acknowledged to be ill-supplied early in the COVID-19 outbreak [8]. Lack of such equipment was widely perceived to have contributed to greater disease ingression, greater transmission after introduction and higher mortality and morbidity within the UK care home sector. We used anonymised care home tracker data that reported on both staffing levels and PPE availability for individual care homes in the county of Norfolk, eastern England UK, in early 2020. We explored which identifiable care-home-specific risk factors could be linked to either ingression or spread of COVID-19 after ingression. It was hoped this analysis could help inform future infection control strategies specific to reducing ingression and spread of COVID-19 within elderly care homes.

## METHODS

### Data

This is a secondary analysis of care home capacity tracker data (carehomes.necsu.nhs.uk) that is available to adult social care departments at English county councils. The data generator (North of England Commissioning Support Unit), data provider (Norfolk County Council) and Faculty of Medicine and Health Sciences Ethics Research Ethics Committee at UEA (reference 2019/20-130) approved the research. Author TW extracted the information for Norfolk, removed true identifiers and added pseudonymised identifier codes. The pseudonymised data included all operational care homes within county boundaries during the monitoring period. Norfolk is a predominantly rural and coastal county in Eastern England, UK, that extends roughly 55 by 40 miles and has a population of approximately 906,000. Residents of Norfolk are relatively ‘old’ within the UK, with a median age around 45 years which compares to a median age of 40.2 years for all UK residents in mid-2018 [10]. The county is neither especially affluent or deprived but does have areas among the 10% most and least deprived areas in England [11]. Norfolk residents are not ethnically diverse; 96.5% of residents in the 2011 census self-identified as White British or White Other [11]. The percentage of ethnic minorities is even lower among persons age 60+. At this point of writing (end May 2020), death rates in Norfolk have been relatively lower than for rest of England [12, 13].

The data describe infection prevalence from COVID-19 in 307 care homes in Norfolk in the 30 days following 5th April 2020. The purpose of CapacityTracker was to monitor shortages of PPE so PPE information was complete (not missing on any dates). PPE provision was reported in one of three categories (Red, Amber, Green) depending on the availability of each of five types of items: aprons, eye protection, gloves, masks and hand sanitiser on that date.

The coding was converted to an ordinal numeric score with green coded as 1, amber 2 and red as 3. Hence, higher scores indicated progressive decrease in PPE supplies. This allowed us to investigate the impacts of variation in individual components of PPE as well as creating an overall score for ‘PPE problems.’ There was a maximum score of 3 for severe deficiency in any single PPE category, and a maximum combined PPE score of 15 which meant severe (red) shortage of all types of PPE. Other data recorded (daily) were concurrent number of residents who concurrently were suspected or known to have COVID-19, counts of specific types of staff on payroll at the facility, bed capacity or occupancy. Care home managers were instructed to input “the number of residents you consider to be suffering from COVID-19 (including whether or not they have been tested)” [14]. Bed capacity was mostly incomplete (not reported on any date for many of the homes). We therefore did not try to use bed capacity indicators in the models.

Staffing and case counts were reported completely for most homes on most dates. Staff were described in three categories: nurses, care-workers and non-care staff. Nurses and care-workers are both involved with personal and physical care but distinguished by different responsibilities, levels of training and pay grades. Non-care workers include cooks, maintenance, administrative and other employees who don’t normally provide face to face care. Staff were further distinguished as actively available for work, absent (due to leave, sickness and vacancies), or additional (extra staff required to meet the needs of residents during the coronavirus pandemic). The counts of each type of staff were input to the model as one of four count categories: 0-10, 11-20, 21-30 or 31 and over (31+) staff in each of the nurse/care worker/non-care worker categories. Data on some dates were incomplete (staff counts not given on every date for every home) so we used the staff counts on the date with most staff (thus ignoring entries where staff counts were stated as all zeros or dates when only one staff category was reported). This was reasonable because, given the relatively short monitoring period (30 days), the total count of staff positions was unlikely to change. Subsequently, we tested whether case counts on any particular day could be linked to PPE status and/or counts in each staffing category. Any care homes that had no staff counts on any dates were excluded because this meant that the information had never been reported rather than the true counts of staff were zero in all staff categories on all dates.

### Analysis

We used a two-step modelling approach to assess the extent to which cases of COVID-19 were associated with the employed number of staff in the care home broken down by category (care, nurse and non-care worker) and the availability of PPE on presence and rate of spread of disease. The first step was to find correlates with any COVID-19 cases in each home, and the second step was to investigate which factors were linked with onwards spread. We considered but rejected logistic regression for the first step because of strong under-dispersion (high percentage of homes without recorded cases). Logistic regression in presence of such high under-dispersion would lead to a strong under-estimation of variable significance which we viewed as unhelpful for informing infection control policy. Instead, to investigate correlates with disease introduction to homes we used a survival analysis to identify any factors that could be linked to timing of disease introduction. For the survival analyses, we used time to infection (measured in days after 5 April) as a measure of the survival time of care homes being free of disease. We analysed time to infection using Cox proportional hazards models with candidate predictors that were counts in categories of care home employee and PPE scores in either single PPE categories or total combined PPE score. We used a stepwise reduction procedure where the first model included all variables and, followed by removal of non-significant variables from the model. Secondly, we used we used mixed effect modelling [15] to analyse within care home spread following ingression. In the mixed effect models, care home was defined as a random effect to investigate the extent to which the care home employee counts and/or PPE parameters might affect spread of established disease (change in the number of cases in the home through time). We had considered using mixed effect log-linear models to investigate worker numbers and PPE status on each day after adjusting for time since the first case. However, the data were over-dispersed and showed substantial aggregation, so it was preferable to use a generalised mixed effect model with a negative binomial error structure [16]. This strategy accounted for aggregation in cases, and let us extend the models to include a correlation structure that allowed for the repeated measures associated with repeated sampling. The best final models, at either ingression or spread stage, were chosen by keeping only predictors that were significant at p ≤ 0.05. Models were fit in the MASS, Survival and lme4 packages in R [17].

## RESULTS

The supplied dataset comprised 307 care homes in Norfolk, UK. Of these, 30 had incursion by COVID-19 during in the period 5 April - 6 May 2020. 59 homes were removed from the analyses as they had not supplied counts of employees in any category on any date; five of these 59 had reported any cases (total of 14 cases at peak). The useable dataset was therefore 248 Norfolk care homes of which 25 had had any COVID-19 cases (total of 133 cases at end of monitoring period).

### Predictors of disease incursion

Only about 10 percent of the care homes were subject to infection by COVID-19, indicating that the data were under-dispersed. A generalised linear model with a quasibinomial error structure to adjust for the under-dispersion demonstrated that risk of any infection (dichotomous outcome) was significantly related to the number of number of non-care workers (t=4.382, p < 0.001) employed in each establishment. Table 1 shows hazard ratios with 95% confidence intervals and significance levels as p-values for the survival analysis (stage 1 model for any ingression of COVID-19 to care homes). In the final ingression model, the number of non-care workers at the care home was the only significant predictor.

**Table 1.** Survival analysis, hazard ratios with 95% confidence intervals and significance levels

| Variable | Hazard Ratio | (95% CI) | p-values |
| --- | --- | --- | --- |
| Non-care workers employed: |  |  |  |
| ≤ 10 | 1.0 (ref) | -- | -- |
| 11-20 | 6.502 | 2.61-16.17 | < 0.001 |
| 21-30 | 9.870 | 3.22-30.22 | < 0.001 |
| ≥ 31 | 18.927 | 2.36-151.90 | 0.006 |
| Model fit metrics: Concordance= 0.742, standard error = 0.048, Likelihood ratio test= 25.33 on 3 degrees of freedom (df; $p < 0.001$), Wald test = 24.38 on 3 df, $p < 0.001$. | | | |

Timing to infection was significantly related to the number of non-care workers employed (Figure 1). Risk of infection was 6.502 times higher (CI: 2.614 −16.17) in care homes that employed 11 to 20 non-care workers; 9.870 times higher (CI: 3.224 −30.22) in homes employing 21-30 care workers and 18.927 times higher (CI 2.358 :151.90) times higher in care homes employing more than 30 non-care workers

**Figure 1.**
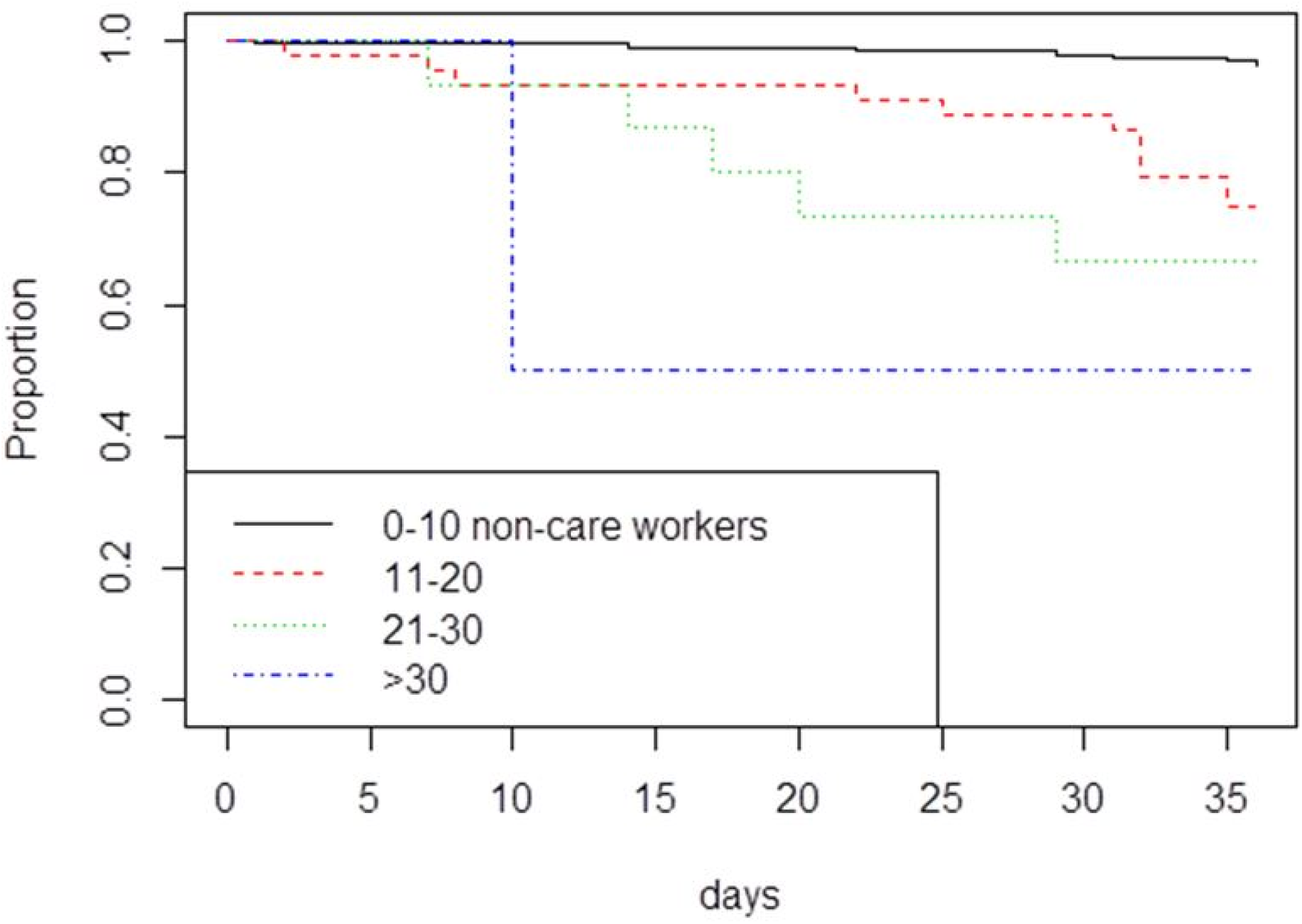
Survivorship curves investigating impacts of numbers of non-care workers employed in care homes on the risk of the care home suffering incursion by COVID-19. Incursion was more rapid in care homes employing more non-care workers.

### Predictors of spread

The exponents of the regression parameters estimates for the best model are shown in Table 2 along with the 95% confidence intervals. The data represent incremental risk (cases) in relation to the increment for each factor. The time increment is expressed in days since beginning of the study (April 5^th^ 2020) as the epidemic proceeded. The coefficients for PPE represent the increments in cases as the state of availability of PPE of different types moved from green through amber to red. It is clear that absence of masks and eye protection had the biggest impact on cases followed by the numbers of care workers. The employee increment is between categories, from 0-10, 11-20, 21-30 or 31+ employees in each group.

**Table 2.**
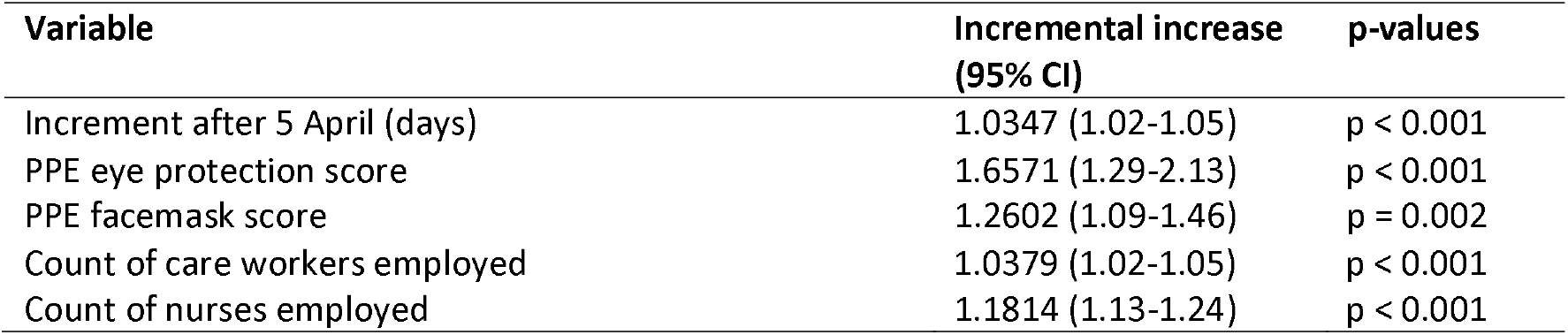
Regression diagnostics for the mixed effect model analysis in factors linked to spread of COVID-19 in care homes. Data represent the incremental increase in cases per unit of the predictor variable.

The daily increment in cases (ie spread) was 1.04. Reduced availability of PPE for eye protection and PPE for facemasks had the greatest impact on spread with coefficients increasing case load by respectively 1.66 and 1.26 per increment (both p < 0.001) on top of staff counts and daily increment effects. Spread (case count increments) also increased with higher staff levels.

The temporal patterns of spread in the 25 care homes and the predicted numbers from the final model are shown in Figure 2. Individual care homes are indicated by anonymised titles (eg., N-00287). There is a reasonably close approximation (visually) between observed and predicted cases in the 25 care homes where disease was recorded and subsequently spread.

**Figure 2.**
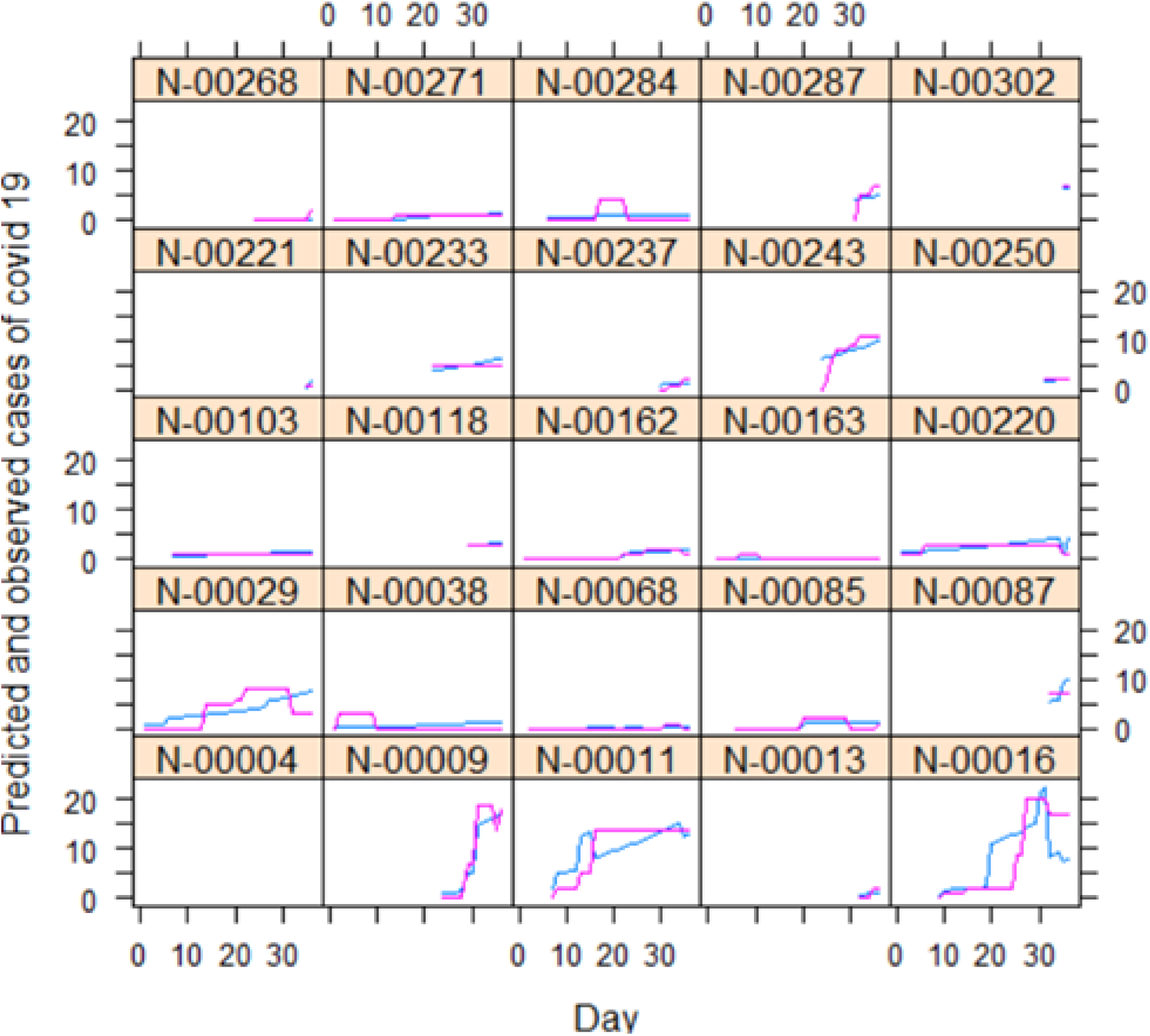
Predicted (blue line) and observed (pink line) numbers of COVID-19 cases in 25 care homes during April-May 2020. Horizontal axis = count of days after 5 April. Model form is mixed effect negative binomial. For predictors see text.

## DISCUSSION

We have shown that entry of COVID 19 into a care home was primarily associated with the number of people not directly involved in the care of residents. We have also shown that once introduced into the home the subsequent spread of COVID-19 was largely associated with inadequate access to PPE, most especially facemasks, and the number of resident-contact workers employed. These findings are not surprising. That the non-care worker category was important may indirectly correspond with low use of PPE among these employees, which meant they had more likelihood of passing the infection to other staff or during brief time spent near residents. Alternatively, non-care workers may be especially likely to work part time and possibly work across several locations. Our dataset did not indicate counts of agency workers (who move between multiple care homes) so we were unable to consider if working in multiple settings was directly relevant. The increased spread subsequent to introduction of COVID19 into a home with inadequate access to PPE is equally unsurprising. Increased case counts linked to higher numbers of care-workers may relate both to inadequate PPE and increased contact rates.

That care homes are particularly vulnerable to the rapid spread of infectious diseases has been described for outbreaks of other infectious diseases, especially norovirus [18], influenza and other respiratory infections [19]. In contrast to outbreaks in health care settings, outbreaks in care homes are relatively less well researched. Twelve years ago Fell [20] investigated the preparedness of care homes for the oncoming Influenza pandemic and concluded that at the time care homes were inadequately prepared for coping with a pandemic. Four factors have been identified [21] that facilitate spread of norovirus in care homes: “missing the diagnosis, care service under pressure, delay in outbreak control measures and patient/resident location and proximity”. Each of these themes has clear resonance with issues around the management of COVID-19 outbreaks in care homes.

Although our research clearly indicated the importance of PPE to reduce disease spread, we argue here that infection prevention and reduction needs to be more multi-faceted than simply supplying adequate PPE and training to use it. Investigation of the Kirklands long-term care facility [9] outbreak highlighted many other problems that can exacerbate a COVID-19 outbreak in this type of setting. It is apparent that better data need to be collected to directly understand better how this sector is vulnerable to an emerging disease like COVID-19. Good understanding of the economic drivers that may compel staff to work in multiple facilities or while symptomatic could help to inform development of different employment policies and practices that might reduce the risks of disease introduction facilitated by common current work patterns. Understanding of how existing work patterns interact with COVID-19 spread is still developing and may be somewhat specific to individual countries [22]. Low index of suspicion was also cited in the Kirklands outbreak, not least because it was difficult to clearly identify affected residents. Persistent coughing and fever are the most diagnostic symptoms for COVID-19 patients, but severity of symptoms is highly variable, even among the elderly [2]. Moreover, care home residents often have respiratory problems [23], especially coughs due to chronic conditions [such as COPD; 24] and/or high susceptibility to minor self-limiting respiratory infections [25]. Regular testing with fast results will be key to help distinguish other cough-inducing conditions from COVID-19 [26].

### Limitations

It is interesting that the data for infection were under-dispersed whilst those for spread were over-dispersed. The former outcome (whether or not a care home had any COVID-19) provides a good justification for using the Cox-proportional hazards model since infection was clearly not a straightforward binomial event; many more events should have occurred for this error structure to have been appropriate for the data. However, in reality, there is likely to have been a spatial component to the existence of disease in the wider community that would have meant that the binomial error model would have been inappropriate without consideration of spatial variation in community. Spatial and social network data interaction between homes were not available to us but would strengthen any future modelling efforts.

Lack of ethnic diversity in Norfolk meant we could not consider whether minority ethnic composition was a factor in disease spread or severity; ethnic diversity seems to be important to disease outcomes among affected care homes in other localities [5]. Considerable efforts have been undertaken to increase supplies of PPE to UK care homes since these April 2020 data were collected [27]. Improvements in procurement processes and supply chains may have changed the balance of future risk factors from what we see in these April 2020 data.

## CONCLUSIONS

Ingression of COVID-19 to Norfolk care homes in April 2020 was most strongly linked to the counts of employed staff in the non-care worker category. Specific detailed research should follow to examine the interaction patterns of all types of staff with each other and with patients. Understanding how often care home staff work in multiple institutions taking on which types of roles with what kinds of physical contact with patients and other staff may be key to improving infection control during a pandemic situation like COVID-19 has created. After disease was introduced, our models implicated lack of eye protection and face masks as the most important risk factors in COVID-19 spread. This information may be helpful for prioritising PPE procurement in future, at least with regard to this respiratory disease. It is worthwhile reiterating that residential care for the elderly is a generally underfunded sector with low pay conditions where staff training has historically been under-valued; ameliorating this situation will not be quick.

## Data Availability

The data are sensitive and we currently don't have permission from the provider to share the data widely.

## Acknowledgements

We appreciate Neil Stevenson of the CapacityTracker service (managed by NHS North of England Commissioning Support Unit) giving consent for the data to be used for research purposes, and Pete Best of the Norfolk and Norwich University Hospitals Trust for facilitating local collaboration between NHS groups and UEA academics.

